# Data-driven forecasting of Flu, RSV, and COVID-19 related outcomes in the United States and Canada via Hankel dynamic mode decomposition

**DOI:** 10.1101/2025.11.12.25339917

**Authors:** William T. Redman, Luke C. Mullany

**Affiliations:** Intelligent Systems Center, Johns Hopkins Applied Physics Lab; Electrical and Computer Engineering Department, Johns Hopkins University

## Abstract

The (large) season-to-season variability and limited dynamical history make the forecasting of infectious diseases a challenging problem. Here, we examine the extent to which advances in data-driven dynamical modeling can provide accurate predictions by benchmarking the performance of one such method, Hankel dynamic mode decomposition (DMD), on the 2024-2025 influenza, respiratory syncytial virus (RSV), and COVID-19 seasons in the United States and Canada. Using Hankel-DMD, we generated weekly forecasts that were submitted to the Center for Disease and Control’s (CDC) FluSight Forecast Hub and the University of Guelph’s AI4CastingHub. Across both Hubs, we find that Hankel-DMD can provide high quality forecasts at the beginning and end of the season, but the times in-between suffer from significant overestimation of the season peak. This leads to worse than baseline performance on FluSight Forecast Hub, when submissions are evaluated across the entire season. Despite this overestimation, Hankel-DMD is found to be the best performing model for forecasting influenza, RSV, and COVID-19 in Canada, although only three other models submitted enough forecasts against which to compare. As this was the first year AI4CastingHub was active, this suggests that Hankel-DMD may be especially useful when expertise is lacking for predicting infectious dynamics in new regions. Retrospective analysis using thresholding and extensions to DMD with memory, a recently developed approach for applying DMD to non-stationary dynamical systems, provide significant improvements during the season peaks. The extent to which this can be achieved in a live forecasting setting remains to be seen. Collectively, our results demonstrate that Hankel-DMD is a promising approach for efficient and interpretable forecasting of infectious diseases and highlights a number of remaining methodological challenges which future work should aim to address.

## 1 Introduction

The need for accurate forecasting of infectious diseases has become increasingly clear in the post-COVID-19 world. Predicting the number of hospitalizations due to COVID-19 or other diseases (e.g., RSV and influenza) several weeks in advance can facilitate the allocation of staff and resources (e.g., ventilators), and better inform the public of the risks associated with travel, large-scale events, and social gatherings. Such information is useful even in the absence of a global pandemic.

To connect government agencies, such as the United States (U.S.) Center for Disease and Control (CDC), with modelers, a number of collaborative initiatives (termed “Hubs”) have been created to crowd-source forecasts of cases and hospitalizations associated with different infectious diseases [1–10]. These platforms have advanced the state-of-the-art in infectious disease forecasting and analytics, with some of the best-performing models developed specifically in response to these modeling Hub requests for contributions [11, 12] and showing improved utility through continual innovation across multiple seasons [13, 14].

Data-driven methods have found considerable success in providing high quality forecasts for these Hubs [3], suggesting that general tools from statistical modeling, time-series analysis, and machine learning (ML) may enable methodologies that can be used across a range of infectious diseases. In the era of increasingly large and powerful ML models, one possible approach to advance prediction accuracy is to train massive models in the context of infectious diseases. However, such models require huge amounts of training data to avoid overfitting, which presents a challenge in settings where relevant data (e.g, number of cases and hospitalizations) have only recently been systematically recorded. Attempts to leverage more general data, beyond only cases and hospitalizations, have been met with limited success [14–16]. In addition, increasing computational costs and model complexity can reduce model interpretation and limit the adoption of such methods at the local region level (e.g., state, province, county, district), where computational resources and staff technical knowledge may be limited.

An alternative approach is to leverage light-weight, data-driven models that explicitly capture the temporally dynamic nature of the data [17]. Recent development of powerful dynamical systems theoretic methods [18–21], grounded in Koopman operator theory [22–24], have found success in a broad range of applications, including capturing dynamics of traffic patterns [25], climate indicators [26, 27], and neural activity [21, 28]. Like standard time-series methods (e.g., auto-regressive integrated moving average, ARIMA), Koopman operator theoretic models are efficient to compute and are interpretable. However, unlike standard time-series models, which tend to only capture linear relationships, Koopman operator theoretic models have greater capacity to capture complex, nonlinear features of the data. While attempts to retrospectively apply Koopman operator theoretic models to infectious disease forecasting have shown promise [29–31], much about their performance and potential remains uncharacterized. In addition, there has been no systematic evaluation of the performance of these methods during an on-going infectious disease season.

To assess the potential strengths and weaknesses of Koopman operator theoretic methods, we used Hankel dynamic mode decomposition (DMD) [20, 21] – a popular approach for data-driven dynamical systems modeling – to generate weekly forecasts for U.S. influenza and Canadian influenza, RSV, and COVID-19 indicators (e.g., hospitalizations, percent positive laboratory detections), from November 2024 until May 2025. As part of our collaborative work with the Atlantic Coast Center for Infectious Disease Dynamics and Analytics (ACCIDDA), we submitted our predictions to two live forecasting hubs: CDC’s FluSight Forecast Hub and University of Guelph’s AI4CastingHub. Across all submissions, we find that our Hankel-DMD model performs well during the initial rise and later decay of infectious disease indicators, consistently outperforming baselines and other models. However, our Hankel-DMD models exhibit significant overestimation during the peaks of the season. In the case of the CDC’s FluSight Forecast Hub, these large forecasts led to full-season performance that was worse than baseline model performance. In the case of the University of Guelph’s AI4CastingHub, our Hankel-DMD model ranked as the top performer. Retrospective analysis reveals that relatively simple and sensible mitigation strategies can be incorporated to improve the performance of Hankel-DMD models, including “DMD with memory” [31] a recent extension developed for non-stationary dynamical systems.

We believe our results using Koopman operator theoretic methods to forecast infectious disease data demonstrate the flexible and general nature of these approaches, while additionally highlighting the need to further develop numerical tools that can improve their performance on non-stationary dynamical data.

## 2 Methods and Materials

### 2.1 Koopman Operator Theory

Let **x**(*t* + 1) = *f* [**x**(*t*)] be a discrete-time dynamical system (i.e., *t*∈ ℕ), where **x** ∈ ℝ^*n*^ and *f* : ℝ^*n*^→ ℝ^*n*^ is a dynamical map. In the setting of infectious disease modeling, **x** could correspond to the number of hospitalizations due to influenza across all 50 U.S. states (*n* = 50), and *f* may correspond to the dynamics associated with influenza transmission and symptomatology.

If *f* is linear, then *f* [**x**(*t*)] = *A***x**(*t*) for some *A* ∈ ℝ^*n*×*n*^. Identifying the relevant dynamical features of *f*, as well as forecasting the future behavior of **x**, amounts to studying the eigenvalues and eigenvectors of *A*. In general, however, *f* is not linear, particularly in infectious disease settings. One powerful framework for dealing with this non-linearity is Koopman operator theory [22–24], which takes the perspective of the dynamics occurring not in the state-space [i.e., *x*(*t*)], but instead, in a lifted function space, *ℱ*. More precisely, let *ψ* : ℝ^*n*^ → *ℱ* be a lifting function. For example, *ψ* could be a polynomial (**x**^*p*^, where *p* ∈ ℕ) or trigonometric function [sin(**x***π/m*), where *m* ∈ ℕ]. Koopman operator theory considers the dynamics of Ψ[**x**(*t*)] = **z**(*t*) ∈ *ℱ*, where Ψ = {*ψ*_1_, …, *ψ*_*L*_}. If *ℱ* and Ψ are suitably chosen, then

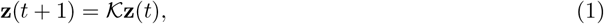

where *𝒦*∈ ℂ^*N* ×*N*^, for some *N > n*, is called the Koopman operator. That is, by lifting to the function space, the dynamics become linear. While in principle, the dimensionality of the function space needed to achieve linearity is infinite (i.e., *N* = ∞), a number of numerical methods have been developed to estimate finite-dimensional approximations of the Koopman operator [19–21, 32], *K*∈ ℂ^*N* ×*N*^ for *N <* ∞, such that **z**(*t* + 1) ≈ *K***z**(*t*).

In the case that a “good” *K* can be found, understanding the relevant dynamical features of *f*, as well as forecasting the future behavior of **x**, amounts to studying the eigenvalues and modes associated with *K*. The state of **z**(*t*), *η* ∈ ℕ steps into the future, can be forecasted as

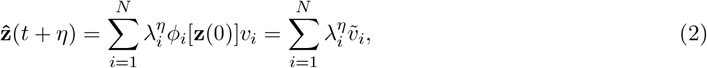

where the triplet (*λ*_*i*_, *ϕ*_*i*_, *v*_*i*_) are the associated Koopman eigenvalues, eigenfunctions, and modes, and 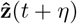 is the forecasted value. For simplicity, we consider 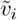, which are the Koopman modes scaled by the Koopman eigenfunctions. That is, 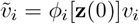. Note that the *ϕ*_*i*_[**z**(0)] are Koopman eigenfunctions, which is dependent on the initial lifted state **z**(0). If the lifted state includes the state-space state, that is **z**(*t*) = [**x**(*t*), …]^⊺^, then the value of 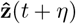 provides information on the value of the forecasted state-space variables, 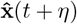.

### 2.2 Hankel–DMD

One popular approach for approximating *K* (Eq. 1) is dynamic mode decomposition (DMD) [33], which has many variants [19–21, 32]. Given a set of lifting functions Ψ and a sequence of *T* ∈ ℕ snapshots of the state-space variables, DMD computes *K* via

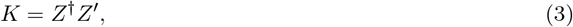

where *Z* = [**z**(1), …, **z**(*T*− 1)] ∈ ℝ^*N* ×*T* −1^, *Z*^′^ = [**z**(2), …, **z**(*T* )] ∈ ℝ^*N* ×*T* −1^, **z**(*t*) = Ψ[**x**(*t*)], and † is the pseudo-inverse.

The original implementation of DMD used the identity lifting function, Ψ(**x**) = **x** [33]. While this can converge to a good approximation of the Koopman operator in specific settings [18], in general this is not sufficient. Extended DMD (EDMD) leverages a dictionary of functions for *ψ* [19], which can be learned via dictionary learning [34]. Similarly, kernels can be used via Kernel DMD [32] to approximate *K*. While these approaches are powerful, they require well-chosen kernels or dictionary functions, and can be computationally expensive. An alternative that has found broad utility is Hankel-DMD [20, 21], which utilizes time-delays as lifting functions. That is,

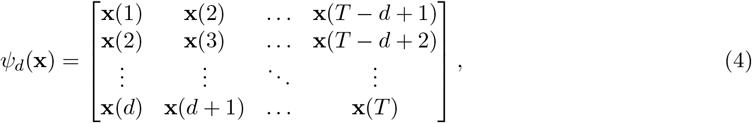

where *d* ∈ ℕ is the number of time-delays. The use of time-delays is a well established practice in dynamical systems theory, as it is known that time-delays can enable reconstruction of dynamical attractors from limited observations [35].

Infectious disease dynamics are generally not only non-linear, but non-stationary, exhibiting large differences in dynamical regimes across time (e.g., across different influenza seasons [12]). Extensions of Koopman operator theory to non-autonomous dynamical systems exist [36–38], with Eq. 2 becoming

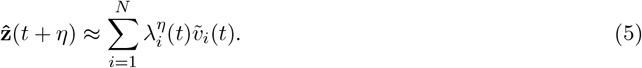

While numerical approaches for computing this time-dependent mode decomposition have been developed [37, 39, 40], their implementation is challenging [37] and they have found limited success when utilized on real-world data. An alternative to learning the representations of Eq. 5 is to instead compute the Koopman mode decomposition not over all *T* snapshots, but on a subset of them. That is, Eq. 3 can be evaluated over temporally-local data *Z*(*t, ω*) = [**z**(*t* − *ω*), …, **z**(*t* − 1)] and *Z*^′^(*t, ω*) = [**z**(*t* − *ω* + 1), …, **z**(*t*)],

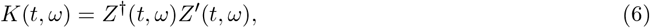

where *ω* ∈ ℕ is called the sliding window. The associated Koopman eigenvalues, *λ*_*i*_(*t, ω*), and modes, 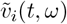, can then be computed via Eq. 5 and used to forecast *η* steps into the future,

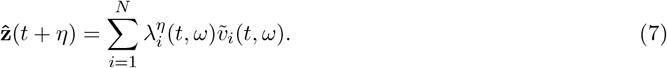

A schematic illustration of Hankel-DMD applied to data from a sliding window is provided in Fig. 1, where first the time-delaying is performed (Fig. 1A), which generates the lifted states (Fig. 1B). These lifted states can be viewed as points in the time-delay coordinate space, which DMD then models (Fig. 1C).

**Figure 1.**
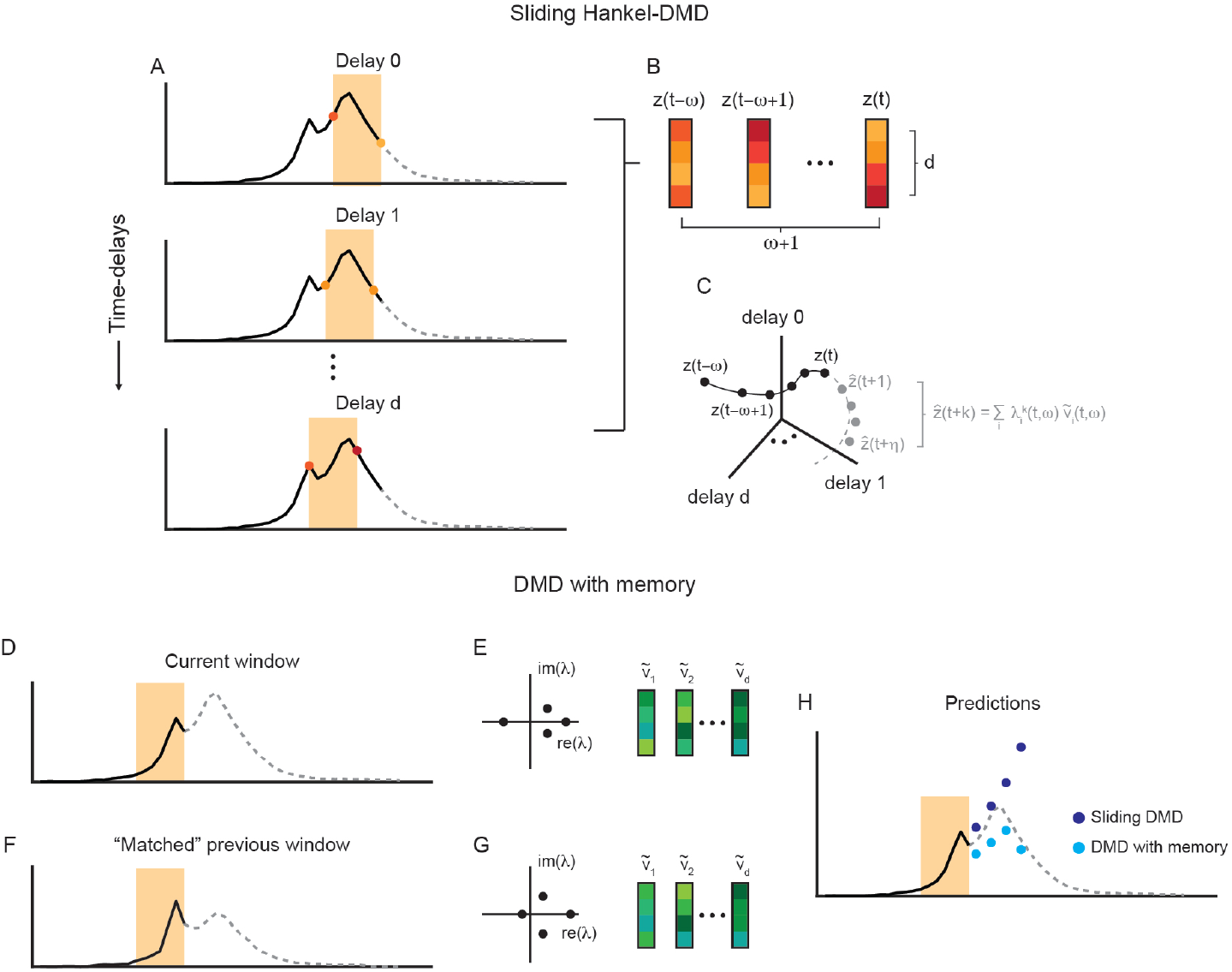
Schematic illustration of sliding DMD and DMD with memory. (A) Given a time-series (known data shown in black, future data shown in gray dashed line), sliding DMD constructs a temporally-local model by time-delaying the data (i.e., shifting the time-series to the left) *d* times and considering only the data within a window of length *ω* (orange shaded rectangle). (B) The time-delaying can be viewed as creating *ω* + 1 vectors of length *d*. (C) These vectors can be viewed as lifted states evolving in the time-delay coordinate space. Sliding DMD learns a model in this space, which can be used to forecast the future states of the system (gray dots). (D)–(H) Instead of relying solely on the sliding DMD model, DMD with memory compares the eigenvalues and modes (E) associated with the current window (D) to all of those associated with previously seen windows. In some cases, there may be a “match”, in which case a window in the past (F) has similar eigenvalues and modes (G). If a match is found, the known information about what happened in the past (F) can be leveraged to make a prediction about the future (H), which may result in better forecasting.

This construction of temporally-local Koopman operator models has found success on complex data sets [25, 30], and can be especially performant when the dynamics are approximately stationary at the time-scale of the sliding window, *ω*. For the remainder of the paper, when referring to sliding DMD, we mean Hankel-DMD applied to a sliding window.

### 2.3 DMD with Memory

Sliding DMD’s effective “forgetting” of past dynamics can prevent the temporally-local Koopman operator, *K*(*t, ω*), from being constructed with data coming from multiple different dynamical regimes, thus avoiding learning a poor approximation [37]. However, if there are re-occurring dynamics – due to repeating latent variables (e.g., similar weather patterns or influenza strains across seasons) – then sliding DMD is not able to leverage this past information to improve its predictions. Qualitative similarities of influenza season dynamics across several years [31], motivates the use of forecasting methods that are able to identify past time-windows with relevant dynamics and use that information for forecasting.

Recently, one such approach was developed, termed “DMD with memory” [31]. This framework has five steps, summarized below:

1. **Computing the Koopman mode decomposition:** The Koopman eigenvalues and modes, *λ*_*i*_(*t, ω*) and 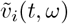, are approximated via DMD.
2. **Comparing current dynamics to past dynamics:** *λ*_*i*_(*t, ω*) and 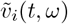 are compared to the Koopman eigenvalues and modes associated with all previously seen time-windows, *λ*(*t*^′^, *ω*)_*i*_ and 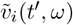, for *t > t*^′^ + *η*. In particular, the distance between Koopman eigenvalues is computed for all pairs via the Wasserstein distance, *d*_*λ*_(*t, t*^′^) = *W*_1_[*λ*(*t, ω*), *λ*(*t*^′^, *ω*)], and the distance between Koopman modes is computed for all pairs via a normalized Euclidean distance, 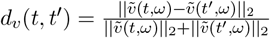.
3. **Identifying dynamical “matches”:** Given two distance thresholds, *ε*_*λ*_, *ε*_*v*_ ∈ ℝ^+^, a dynamical match between the current dynamics and the dynamics that occurred during the time-window *t*^′^ exists if *d*_*λ*_(*t, t*^′^) *< ε*_*λ*_ and *d*_*v*_(*t, t*^′^) *< ε*_*v*_. If more than one match is found, the time-window with the smallest summed distance is chosen. That is, *t*_match_ = min_*t*_*′* {*d*_*λ*_(*t, t*^′^) + *d*_*v*_(*t, t*^′^) | *d*_*λ*_(*t, t*^′^) *< ε*_*λ*_, *d*_*v*_(*t, t*^′^) *< ε*_*v*_}.
4. **Forecasting future dynamics:** If a match exists, then the known dynamics that followed the time-window at *t*_match_ are used for forecasting. In particular, if there is a match, then 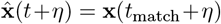. Scaling can be introduced to better align the amplitudes of the forecast. If no match exists, then the standard sliding DMD prediction (Eq. 6) is used.
5. **Saving Koopman eigenvalues and modes:** After forecasting, *λ*_*i*_(*t, ω*) and 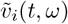 are saved.

More details on each of these steps can be found in Redman et al. (2025) [31]. A schematic illustration of this approach can be seen in Fig. 1. The current sliding window (Fig. 1D) has Koopman eigenvalues and modes (Fig. 1E) that are similar to another sliding that was seen previously (Fig. 1F, G). Therefore, instead of using the sliding DMD forecast, which may not accurately capture the transition between two dynamical regimes (Fig. 1H, dark blue dots), the known dynamics that occurred subsequent to the matched window are used as a forecast (Fig. 1H, light blue dots).

Application of DMD with memory to non-stationary time-series can lead to large improvements in performance. While these results highlight the potential of DMD with memory, the framework as developed by Redman et al. (2025) [31] was limited to utilization of a single “memory” (Step 3), *t*_match_, and a forecasting that is an exact (up to scaling) recall of the past dynamics (Step 4), **x**(*t*_match_ + *η*).

To allow for a more generative process that can better leverage relevant past dynamics, we develop the following two extensions:

1. **Allowing for multiple matches:** Instead of using only a single time-window seen previously, *t*_match_, we consider multiple time-windows with Koopman eigenvalues and modes sufficiently similar to *λ*(*t, ω*) and 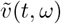.
2. **Forecasting via averaging across matches:** Instead of using exactly what happened previously after time-window *t*_match_, **x**(*t*_match_ + *η*), we average what happened previously after all time-windows that were deemed matches. That is,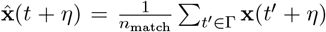, where Γ is the set of time windows in the past that had matched dynamics and *n*_match_ is the number of matches found (i.e., *n*_match_ = |Γ|).

For more details on our implementation of DMD with memory can be found in Appendix A.

### 2.4 FluSight Forecast Hub

The CDC’s FluSight Forecast Hub is an initiative that has been on-going since the 2010-2011 season [3, 4, 6, 10]. During the 2024-2025 season, modeling teams were asked to provide weekly forecasts for the number of hospital admissions due to lab-confirmed influenza at 1-, 2-, and 3-week horizons. Forecasts are in the form of probabilistic quantiles, with predictions for the [1, 2.5, 5, 10, …, 95, 97.5, 99] percentiles. Forecasts are made at both the national and state level. States vary significantly in their population, geography, and size, making this a challenging problem. In addition to the national and state-level targets, the FluSight Forecasting Hub also requests forecast outputs for Washington D.C. and Puerto Rico. In total, there are 53 target regions, three target horizons, and 23 percentile targets. Across the season, 38 models (including our model, with submission name JHUAPL-DMD) submitted a sufficient number of forecasts to be included in a performance evaluation, as determined by the CDC FluSight team [41].

### 2.5 AI4CastingHub

AI4CastingHub is a recent initiative from the University of Guelph to source forecasts for infectious diseases in Canada. The 2024-2025 season is its first active season. Within the AI4CastingHub, there are two datasets: Forecasting Hospital Bed Occupancy Data for Ontario Respiratory Virus Activity (Ontario dataset) and Respiratory Virus Detection Surveillance System (RVDSS dataset). For sake of brevity, we focus in this work on our RVDSS datset forecasts, although we additionally made forecasts for the Ontario dataset and found similar results.

For the RVDSS dataset, modeling teams were asked to provide weekly forecasts for the percent of laboratory detections across Canada that were positive for RSV, COVID-19, and the influenza at 1-, 2-, and 3-week horizons. Forecasts were submitted in the form of probabilistic quantiles, with predictions for the [2.5, 10, 25, 50, 75, 90, 97.5] percentiles, and were made at the national and the province/territory level. As in the US, these sub-national administratve units vary substantially in population, geography, and size. In addition to the national and province/territory level targets, there are also three collections of provinces/territories (Atlantic, Prairies, and Territories) which were further aggregated into additional targets. In total, there are 17 target regions, three target horizons, and seven target percentiles. Across the season, four models (including our model, with submission name JHUAPL-DMD) submitted sufficient weekly forecasts to be evaluated.

### 2.6 Quantile Forecasting

Given a series of snapshots of state-space variables, {**x**(1), …, **x**(*T* ) }, and a set of lifting functions, Ψ, sliding DMD is deterministic. To generate quantiled forecasts, as is required for AI4CastingHub and FluSight Forecast Hub submissions, we perturb the sequence of **x**(*t*) by adding sampled noise,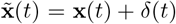. Noise is generated by sampling from Gaussian processes, *δ*_*i*_(*t*) ∼: *𝒩* (0, *σ*_*i*_), with *σ*_*i*_ being fit on each target’s distribution of differences in consecutive snapshots, *x*_*i*_(*t*) − *x*_*i*_(*t* − 1) (see Appendix B). To obtain sufficient statistics, *n*_noise_ ∈ ℕ realizations of noise are sampled, leading to *n*_noise_ perturbed sequences, 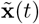. For each sequence, sliding DMD is used to generate forecasts and from the distribution of forecasts, for each target location and horizon, the percentiles are computed.

### 2.7 Evaluation Metrics

A natural way to assess the performance of each model’s forecast is to compute the median average error (MAE) of the predicted 50^th^ percentile and the observed data. However, this approach does not take into account the other forecasted quantiles. To quantify the performance of an individual model’s forecast, for a given quantile *α*, the interval score (IS) can be used [42],

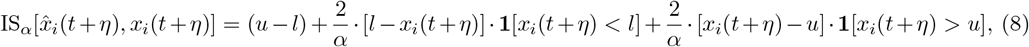

where *α* ∈ *A* = {*α*_0_, …, *α*_*P*_ } is a given quantile, 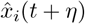 is the forecast for the *i*^th^ region, *x*_*i*_(*t* + *η*) is the observed value, **1**(·) is the indicator function, 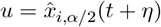 is the forecasted value at the *α/*2 quantile, and 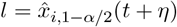 is the forecasted value at the 1 − *α/*2 quantile.

Eq. 8 is for a single quantile. To quantify the extent to which the overall forecasted quantiles match the observed data, the weighted IS (WIS) can be computed [42],

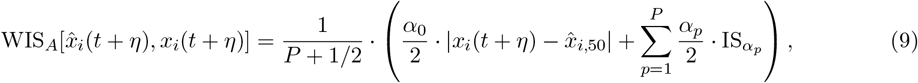

where 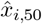 is the median forecast for the *i*^th^ region.

The computed WIS and MAE are publicly available by AI4Casting Hub^1^, and the computed WIS and MAE are publicly available by FluSight Forecast Hub^2^. We note that, while AI4CastingHub reports the median value across all submissions, FluSight Forecasting Hub reports the mean. We align our reporting of metrics to the conventions of the two Hubs.

## 3 Results

### 3.1 FluSight Data Forecasting

The U.S. 2024-2025 influenza season was the third full season that the number of hospitalizations attributed to influenza was used as the forecasting target for FluSight Forecasting Hub. Unlike the previous two seasons, the 2024-2025 season exhibited: 1) a delayed rise in hospitalizations (peak week being February 8, 2025, as compared to December 5, 2022 and December 30, 2023); 2) a greater peak magnitude (*>* 50, 000 weekly hospitalizations in 2024-2025, as compared to *<* 30, 000 weekly hospitalizations in 2022-2023 and *<* 25, 000 in 2023-2024); 3) two peaks, separated by several weeks (Fig. 2A) [10, 41, 43].

**Figure 2.**
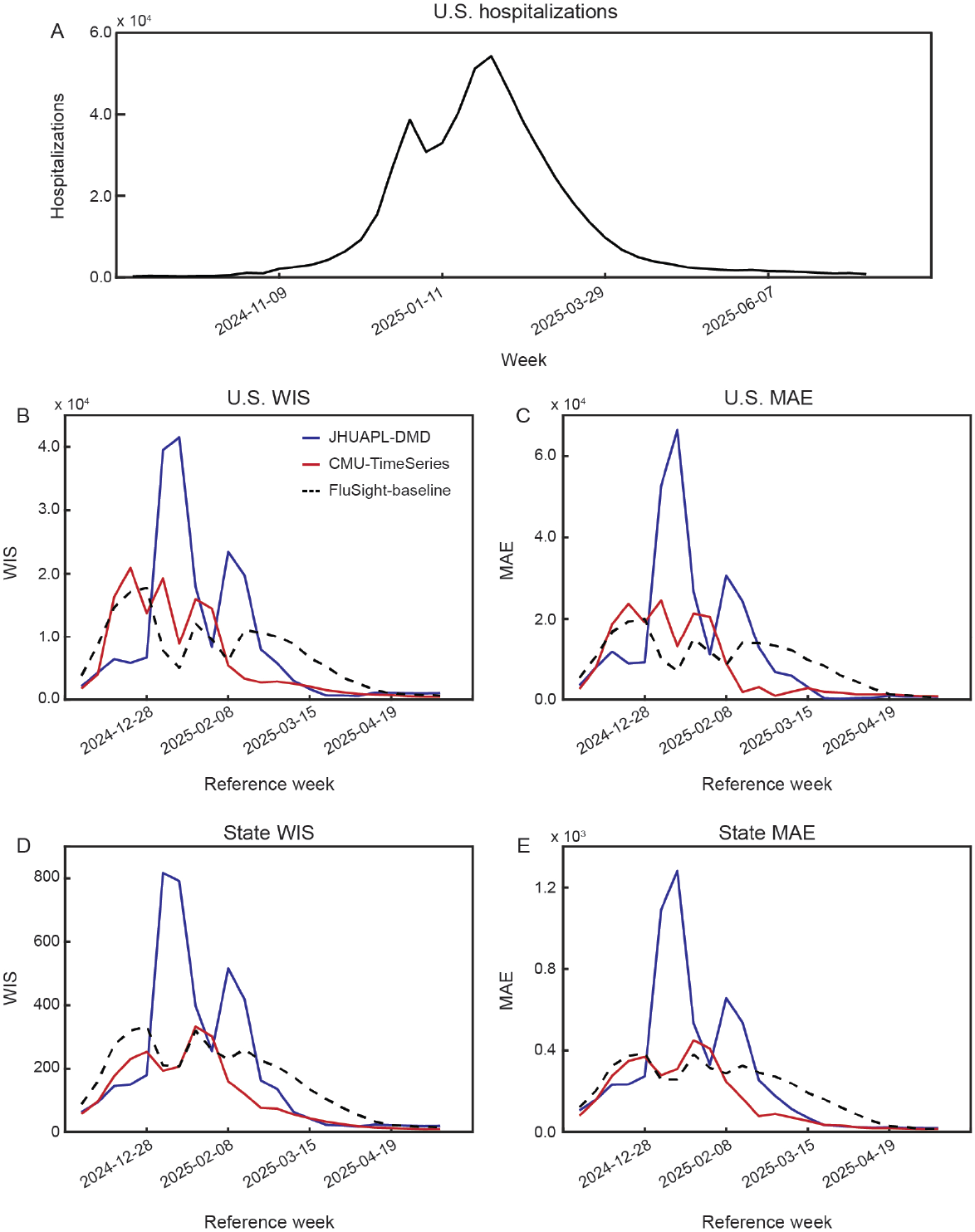
Performance of JHUAPL-DMD model forecasting U.S. influenza hospitalizations, by submission week. (A) Total number of influenza hospitalizations in the U.S. over the 2024-2025 forecast season. (B)–(C) National WIS and MAE, across submission weeks, for JHUAPL-DMD, CMU-TimeSeries, and FluSight-baseline. (D)–(E) State WIS and MAE, across submission weeks, for JHUAPL-DMD, CMU-TimeSeries, and FluSight-baseline.

During the initial five weeks of the season, we find that our JHUAPL-DMD model outperforms not only the FluSight-baseline model, which predicts that the number of hospitalizations will remain fixed at the current value, but also the CMU-TimeSeries model, historically one of the best performing FluSight models. This is true both at the national and state level, for WIS and MAE (Fig. 2B–E – reference week up to 2024-12-28). Examining individual forecast trajectories during this early period reveals that the sliding DMD model predicts continually increasing hospitalizations, well matching the true dynamics (Fig. 3, top row – blue lines). This comes from the fact that the sliding DMD model is only given temporally-local data, across which there is a general increase. In contrast, the forecasts of the CMU-TimeSeries model predict a more modest increase, followed by a decrease in hospitalizations (Fig. 3, top row – red lines). This is likely due to the leveraging of information from the past seasons, which exhibited earlier peaks of hospitalization, at lower magnitude.

**Figure 3.**
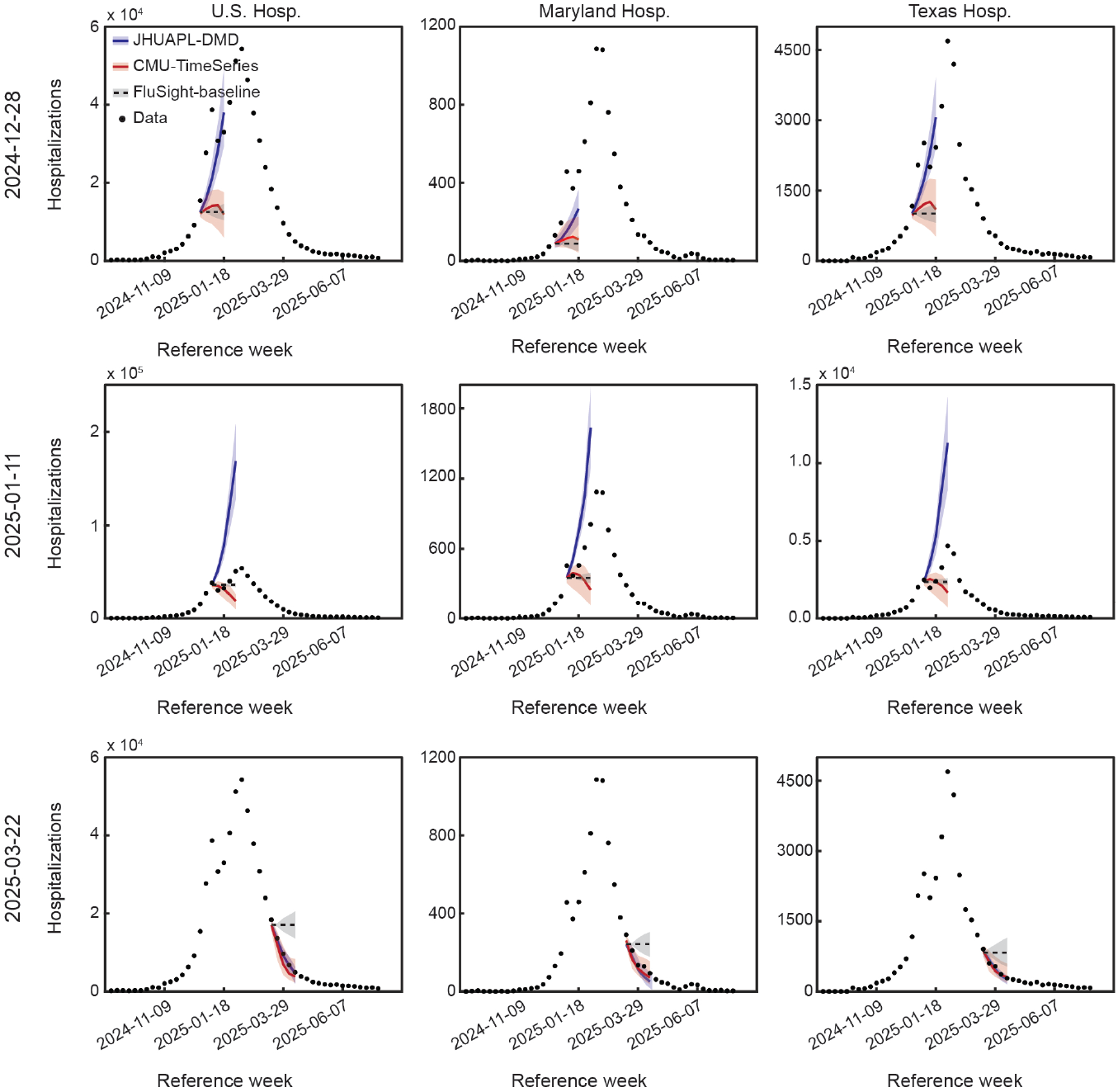
Example U.S. influenza hospitalization forecast trajectories. Submitted forecasts for the U.S., Maryland, and Texas influenza hospitalizations by JHUAPL-DMD, CMU-TimeSeries, and FluSight-baseline models. Example submission weeks were picked for being at the beginning (top row), during the first peak (middle row), and at the end (bottom row) of the influenza season.

Similar to the beginning of the season, the JHUAPL-DMD model outperforms the FluSight-baseline model and performs comparably to the CMU-TimeSeries model at the end of the season (Fig. 2B–D – reference week from 2025-03-01). Examining individual forecast trajectories during this late period demonstrates that the sliding DMD model is able to capture the exponential-like decay of the hospitalizations (Fig. 3, bottom row – blue lines).

However, during the peak part of the season, the JHUAPL-DMD model performs significantly worse than both the CMU-TimeSeries and the FluSight-baseline models (Fig. 2B–D – references week from 2025-01-04 to 2025-02-22). Examining individual forecast trajectories during the peak period demonstrates that this poor performance is due to overestimation of peak hospitalization magnitude (Fig. 3, middle row – blue lines). These very large forecasts lead to significant MAE and WIS errors, which dominate when evaluating the mean performance across the season (Table 1 – JHUAPL-DMD).

**Table 1.**
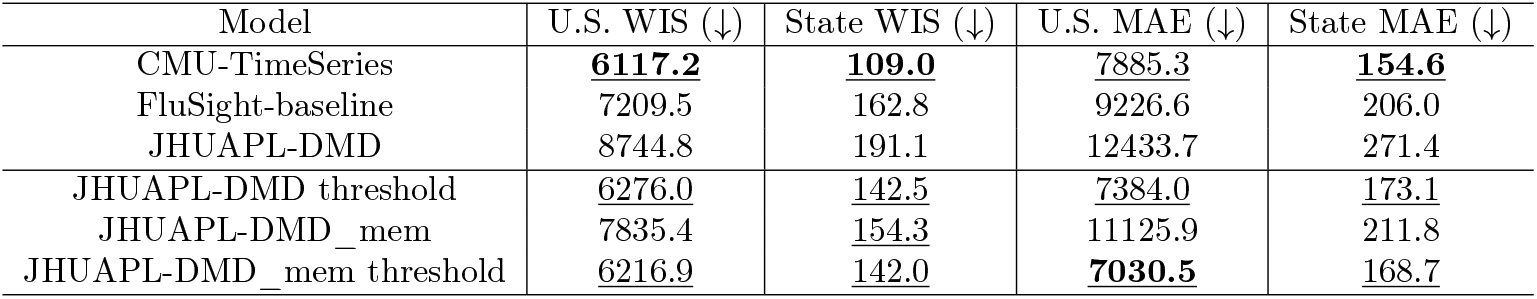
National and state performance of CMU-TimeSeries, FluSight-baseline, and JHUAPL-DMD models across 2024-2025 U.S. influenza season. Model with best performance in each column is bolded. All models with performance better than the FluSight-baseline model are underlined.

| Model | U.S. WIS (↓) | State WIS (↓) | U.S. MAE (↓) | State MAE (↓) |
| --- | --- | --- | --- | --- |
| CMU-TimeSeries | **6117.2** | **109.0** | <u>7885.3</u> | **154.6** |
| FluSight-baseline | 7209.5 | 162.8 | 9226.6 | 206.0 |
| JHUAPL-DMD | 8744.8 | 191.1 | 12433.7 | 271.4 |
| JHUAPL-DMD threshold | <u>6276.0</u> | <u>142.5</u> | <u>7384.0</u> | <u>173.1</u> |
| JHUAPL-DMD_mem | 7835.4 | <u>154.3</u> | 11125.9 | 211.8 |
| JHUAPL-DMD_mem threshold | <u>6216.9</u> | <u>142.0</u> | **7030.5** | <u>168.7</u> |

A simple way to attenuate the explosion in predicted hospitalizations is to impose a threshold on the value of each forecast. If we set the threshold to be the maximum number of hospitalizations, per region (state or nation), seen from the previous two U.S. influenza seasons, we find that the performance of the JHUAPL-DMD model is significantly improved, with its MAE and WIS decreasing below the FluSight-baseline and approaching the performance of CMU-TimeSeries (Table 1 – JHUAPL-DMD threshold).

While this demonstrates that, up to overestimating the peaks, the JHUAPL-DMD model is performant and that its performance can be easily improved, such a naïve thresholding approach has several important drawbacks. First, setting the threshold value to the maximum of what has been previously observed is problematic when the current season is more severe than prior seasons. As already noted, the 2024-2025 season had a significantly greater peak hospitalization magnitude, so our threshold based on the past is too low. And second, thresholding forecasts reduces the amount of temporal information that is provided by the model, as forecasts across multiple time-horizons that are greater than the threshold all get collapsed to a single value. This limits the utility of the model, since determining the hospitalizations trend (increasing vs. decreasing) is of importance.

Motivated by these limitations, we examine whether DMD with memory [31] (Sec. 2.3) can help reduce the explosions in prediction. We find that, during the peaks of the influenza season, a model that uses DMD with memory (which we refer to as JHUAPL-DMD_mem) leads to a large reduction in error (Fig. 4A–B– reference weeks between 2025-01-04 and 2025-03-01), relative to sliding DMD. Examining individual forecast trajectories during the peak period demonstrates that the DMD with memory model can not only avoid unreasonably large forecasts, but can also accurately predict the hospitalization dynamics, especially at the state-level (Fig. 4C, middle and bottom rows).

**Figure 4.**
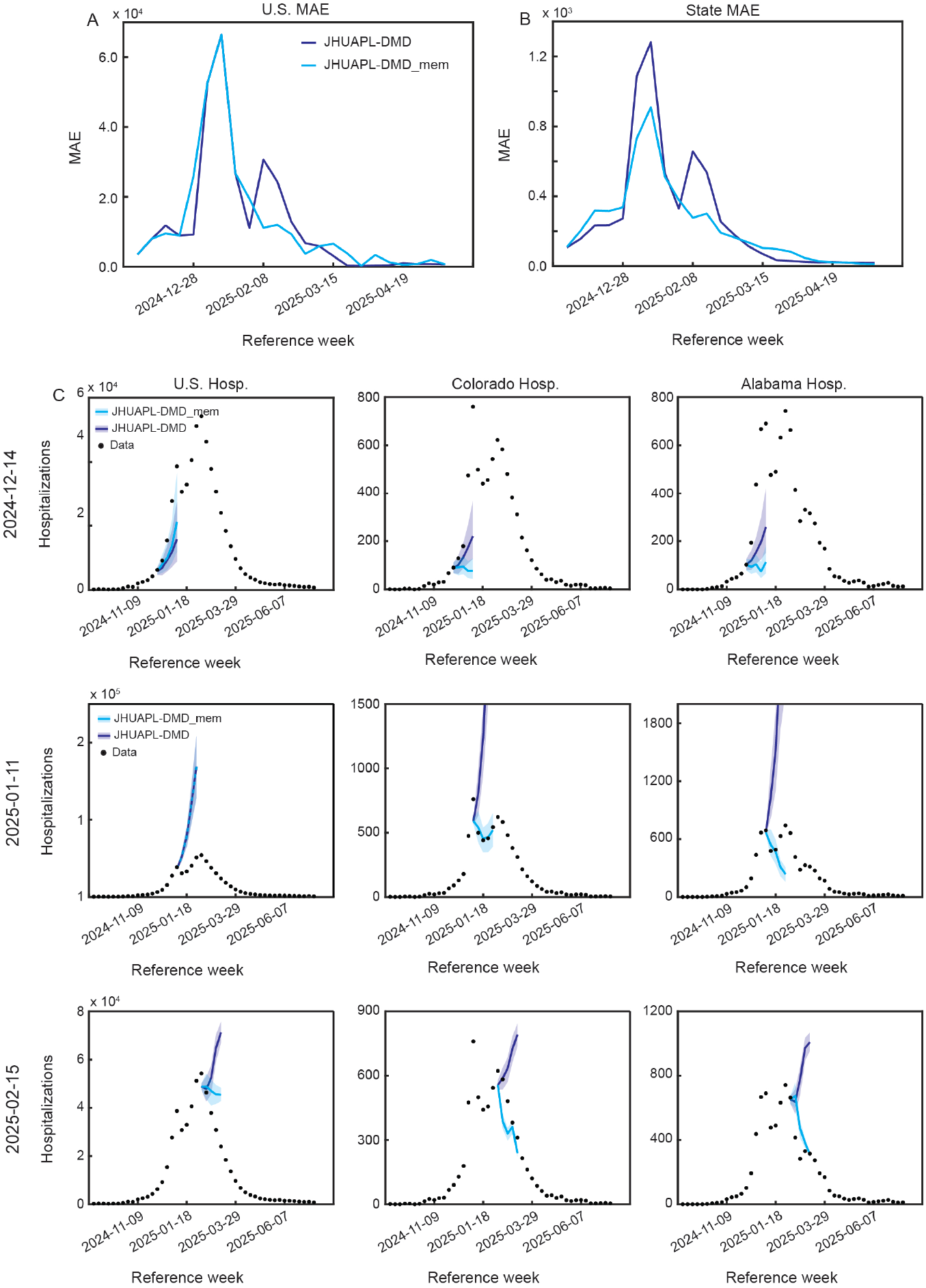
Utilization of memory improves the performance of JHUAPL-DMD during the U.S. influenza season’s peak. (A)–(B) Nation and state MAE, across submission weeks, for JHUAPL-DMD and JHUAPL-DMD_mem. (C) Example JHUAPL-DMD and JHUAPL-DMD_mem forecasts for U.S., Colorado, and Alabama influenza hospitalizations.

While showing promising results during the peak of the season, the DMD with memory model has worse performance, relative to sliding DMD, before the peak (Fig. 4A–B– reference weeks up to 2024-12-28). This is likely due to JHUAPL-DMD_mem prematurely predicting decreases in hospitalization (Fig. 4C, top row), making it similar to the CMU-TimeSeries model forecasts. The DMD with memory model forecasts after the peak have slightly worse performance as well, relative to sliding DMD (Fig. 4A–B– reference weeks after 2025-03-08). These results demonstrate that, while there is a large benefit of using DMD with memory to reduce the overestimation of the predicted peak behavior, it should not be used during the pre- and post-peak weeks, where JHUAPL-DMD already performs well. This can easily be done when performing retrospective analysis, but achieving this in real-time requires more consideration.

Selectively applying the DMD with memory model to forecast the behavior of the peak weeks (and using the sliding DMD to forecast the other weeks) leads to a large improvement in WIS and MAE, at both the national and state level (Table 1 – JHUAPL-DMD_mem). However, because the DMD with memory model makes some of the same forecasts as sliding DMD, in the case that no match to previous dynamics can be found (Sec. 2.3), some of the forecasts may still overestimate the peak hospitalizations. This leads to the mean performance still not being quite as good as the FluSight-baseline model. Therefore, we also threshold the JHUAPL-DMD_mem model forecasts. This improves performance, relative to thresholding without memory (JHUAPL-DMD threshold), bringing the MAE and WIS below that of the FluSight-baseline model (Tab. 1 – JHUAPL-DMD_mem threshold). In addition, the JHUAPL-DMD_mem threshold model has lower national MAE than the CMU-TimeSeries model and gets to within 10% of the CMU-TimeSeries model’s state MAE (Tab. 1).

Taken together, our analysis of the sliding DMD model on the U.S. 2024-2025 influenza season demonstrates both strengths (good performance during the beginning and end of the influenza season) and weaknesses (overestimation of the peak behavior). Using tools designed to better the performance of DMD on non-stationary dynamical systems leads to significant improvement in forecasting the peak of the influenza season. However, the continued need for thresholding suggests that more work is needed to improve the robustness of DMD with memory.

### 3.2 Respiratory Virus Detection Surveillance System (RVDSS) Data Forecasting

Unlike FluSight Forecasting Hub, whose nearly 15 years of activity has enabled modelers to gain extensive experience in U.S. influenza dynamics, the 2024-2025 season was the first season that AI4CastingHub was active. While there are common aspects between the U.S. and Canada, there are several important differences. This includes: 1) the difference in total population (≈ 40 million in Canada; ≈ 335 million in the U.S.); 2) the difference in distribution of population (*>* 38% of the Canadian population lives in the most populous province, Ontario; *>* 11% of the U.S. population lives in the most populous state, California); 3) the difference in population density (maximum and minimum density of 31 and 0.02 persons per km^2^ in Prince Edward Island and Nunavut, respectively; maximum and minimum density of 488 and 0.5 persons per km^2^ in New Jersey and Alaska, respectively); 4) the difference in healthcare (universal public healthcare in Canada; hybrid private insurance and public programs in the U.S.). In addition to these and many other differences, the RVDSS dataset characterizes the percent of positive laboratory detections of influenza, RSV, and COVID-19 over time (Sec. 2.5). This is different from the influenza hospitalizations and influenza-like illness (ILI) targets that the FluSight Forecast Hub have historically used.

Because sliding DMD is data-agnostic, it is a general time-series analysis method. We reasoned that this property would make it especially useful for forecasting the RVDSS data, as it requires little calibration or expert knowledge. We find that, similar to its performance on the U.S. influenza season, the JHUAPL-DMD model performs well at the beginning and end of the Canadian 2024-2025 influenza season, and has large WIS and MAE values during the peak of the season (Fig. 5B–D – blue lines). Note that, due to a bug in our code, two submission weeks, 2024-12-14 and 2025-01-04, incorrectly loaded data and therefore, produced incorrect submissions. For fairness in comparison with other models, we have included these erroneous forecasts in our analysis. This makes interpreting our results at the beginning of the season slightly less straightforward. However, ignoring these two weeks, we see that JHUAPL-DMD has performance relatively similar to the AI4Casting_Hub-Quantile_Baseline (which forecasts the current level of positive laboratory detections). Examining individual forecast trajectories at the Canadian national level, we find that JHUAPL-DMD exhibits similar behavior as it did in forecasting the U.S. influenza hospitalizations (compare Fig. 6, left column with Fig. 3, left column).

**Figure 5.**
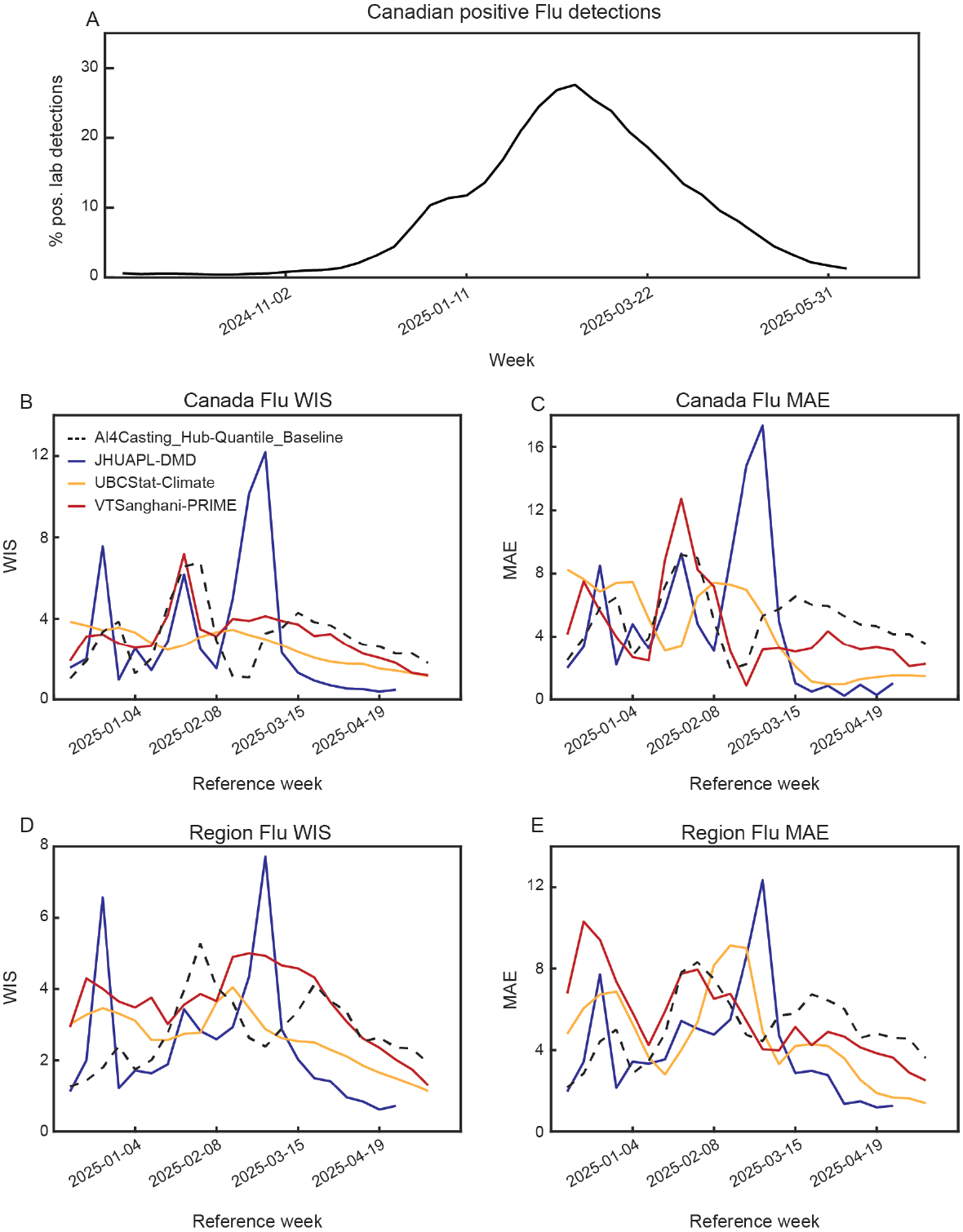
Performance of JHUAPL-DMD model forecasting Canada percentage of positive laboratory influenza detections, by submission week. (A) Percent positive influenza laboratory detections in Canada over the 2024-2025 forecast season. (B)–(C) National WIS and MAE, across submission weeks, for JHUAPL-DMD, UBCStat-Climate, VTSanghani-PRIME, and AI4Casting_Hub-Quantile_Baseline. (D)–(E) State WIS and MAE, across submission weeks, for JHUAPL-DMD, UBCStat-Climate, VTSanghani-PRIME, and AI4Casting_Hub-Quantile_Baseline.

**Figure 6.**
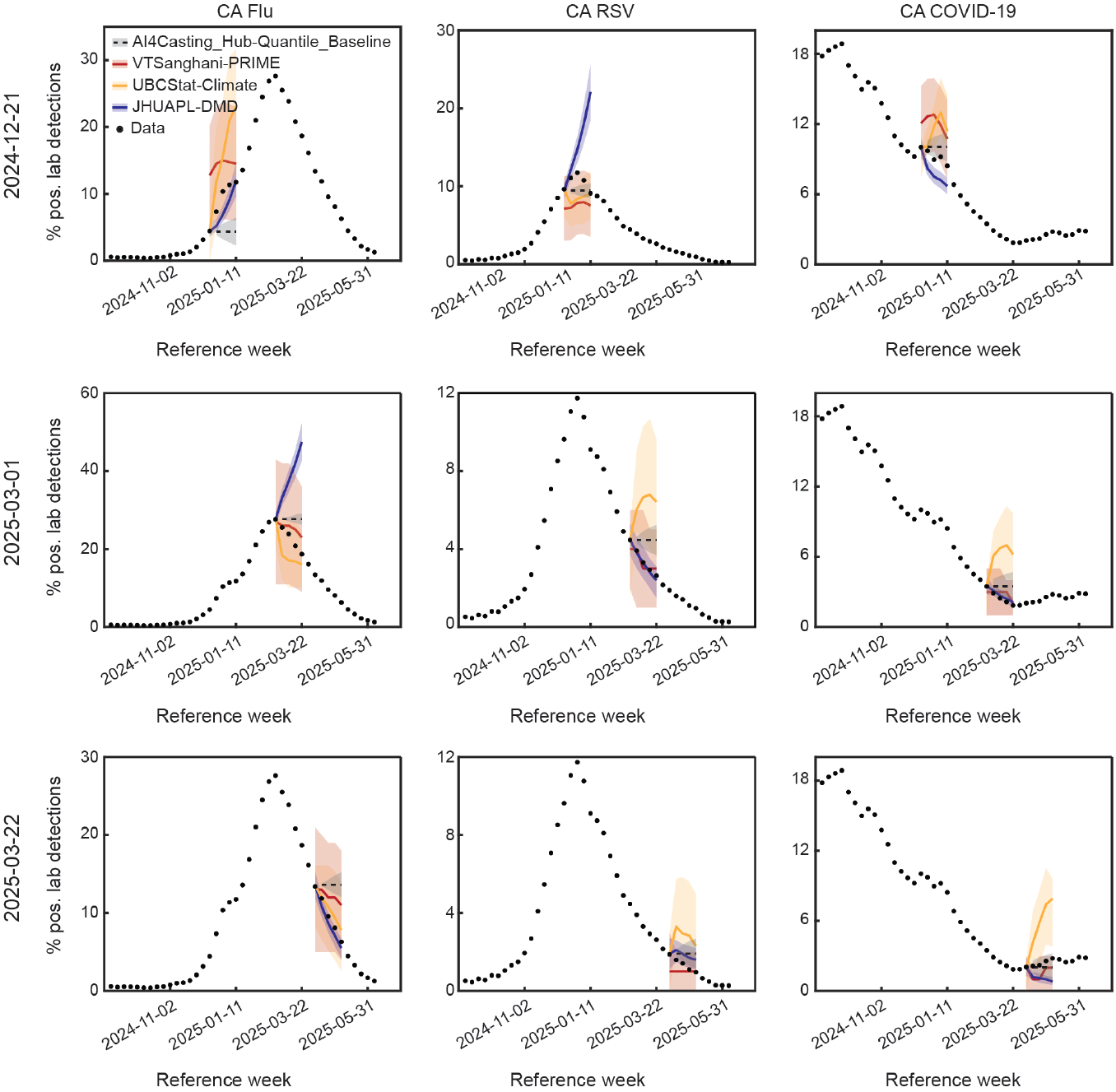
Example Canadian RVDSS influenza, RSV, and COVID-19 percent positive laboratory detections forecasts. Submitted forecasts for percent positive influenza, RSV, and COVID-19 laboratory detections in Canada by JHUAPL-DMD, UBCStat-Climate, VTSanghani-PRIME, and AI4Casting_Hub-Quantile_Baseline models. Example submission weeks were picked for being at the peak of the RSV season (top row), peak of the influenza season (middle row), and beginning of the rise of COVID-19 (bottom row).

We compare the JHUAPL-DMD model forecasts with two other models, VTSanghani-PRIME and UBCStat-Climate, in addition to AI4Casting_Hub-Quantile_Baseline. These were the only models that submitted a sufficient number of forecasts to be evaluated. We find that all three models exhibit large increases in WIS and MAE throughout the season, although the timing of the occurrence of maximum error differs across models (Fig. 5B–D). Evaluating the season-wide performance by taking the median MAE and WIS, we find that the JHUAPL-DMD model has the best performance of all models, at both the national and regional level (Table 2). In addition, the JHUAPL-DMD model’s MAE and WIS are lower than those of the AI4Casting_Hub-Quantile_Baseline model. This is in contrast to the other models. The UBCStat-Climate model does not outperform the baseline on regional WIS and the VTSanghani-PRIME model only outperforms the baseline on national MAE (Table 2).

**Table 2.** National and regional based performance of UBCStat-Climate, VTSanghani-PRIME, AI4Casting_Hub-Quantile_Baseline, and JHUAPL-DMD across 2024-2025 Canadian influenza season. Model with best performance in each column is bolded. All models with performance better than AI4Casting_Hub-Quantile_Baseline are underlined.

| Model | Canada WIS (↓) | Region WIS (↓) | Canada MAE (↓) | Region MAE (↓) |
| --- | --- | --- | --- | --- |
| UBCStat-Climate | <u>2.54</u> | 2.50 | <u>4.02</u> | <u>3.60</u> |
| VTsanghani-PRIME | 2.93 | 3.52 | <u>3.80</u> | 5.34 |
| AI4Casting_Hub-Quantile_Baseline | 2.73 | 2.54 | 4.68 | 4.86 |
| JHUAPL-DMD | **1.50** | **1.72** | **2.38** | **3.18** |

The Canadian 2024-2025 RSV season peaked considerably earlier than the influenza season (compare Fig. 7A with Fig. 5A). Indeed, the peak of the RSV season was a few weeks after the official start of AI4CastingHub RVDSS submissions. This, combined with the erroneous early-season submissions of our model, made evaluating the performance of the JHUAPL-DMD model, at the peak of the season, challenging. However, as can be seen from an example national forecast trajectory during this peak period (Fig. 6, middle column top row), the JHUAPL-DMD model overestimates the peak of the forecast. This suggests a similar story of poor performance at during the peak of the season.

**Figure 7.**
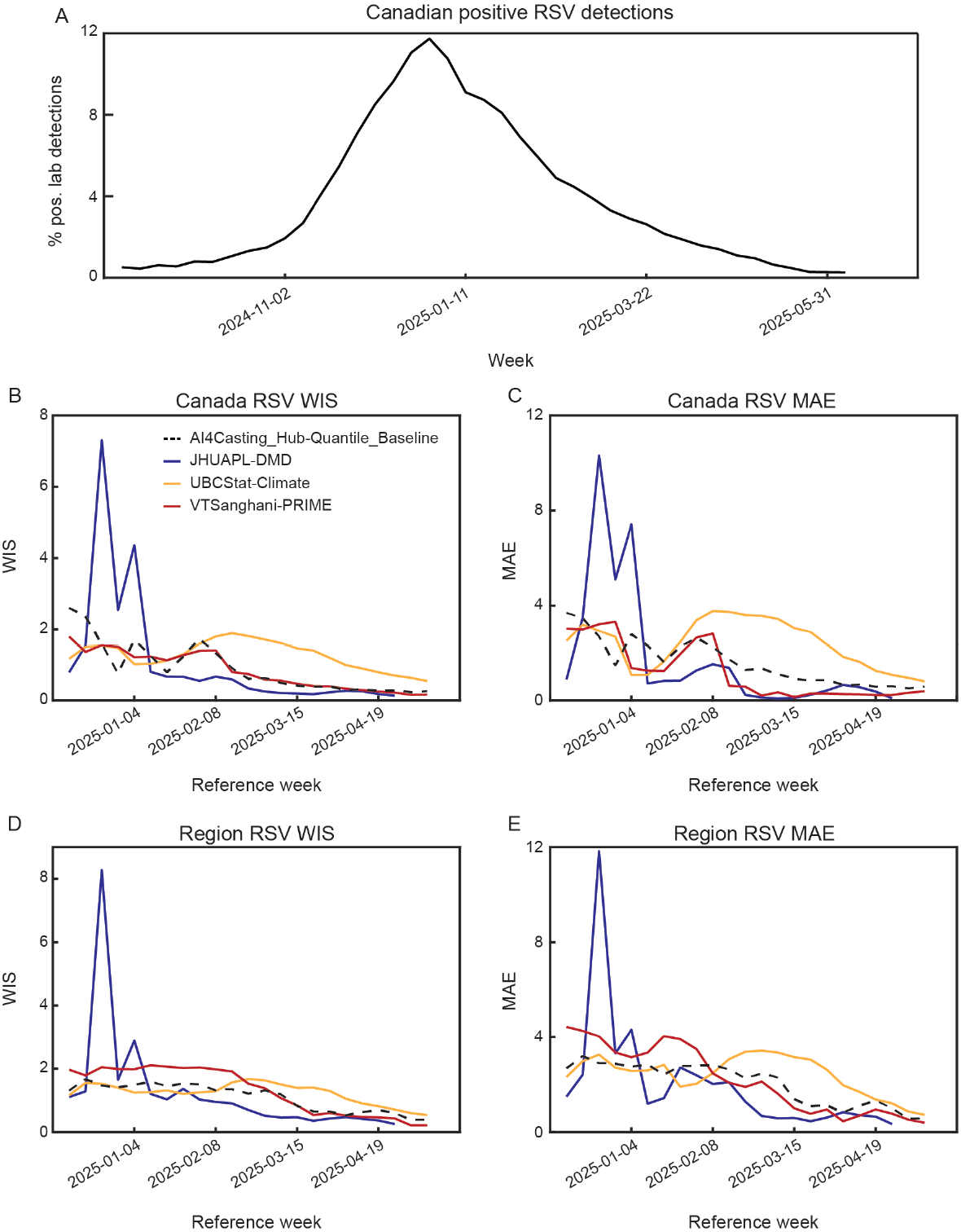
Performance of JHUAPL-DMD model forecasting Canada percentage of positive laboratory RSV detections, by submission week. (A) Percent of positive RSV laboratory detections in Canada over the 2024-2025 forecast season. (B)–(C) National WIS and MAE, across submission weeks, for JHUAPL-DMD, UBCStat-Climate, VTSanghani-PRIME, and AI4Casting_Hub-Quantile_Baseline. (D)–(E) State WIS and MAE, across submission weeks, for JHUAPL-DMD, UBCStat-Climate, VTSanghani-PRIME, and AI4Casting_Hub-Quantile_Baseline.

As found in the U.S. and Canadian influenza forecasts, the JHUAPL-DMD model outperforms all other models for the majority of the post-peak weeks (Fig. 7B–D – reference weeks 2025-01-11 and after). This appears to be due to the fact that, during the later part of the RSV season, the JHUAPL-DMD model consistently forecasts a decrease in positive RSV detections, whereas the UBCStat-Climate model predicts increases and the VTSanghani-PRIME model predicts periods of stable positive RSV detections (Fig. 6, middle column, middle and bottom rows).

Consistent with these observations, we find that the median MAE and WIS, computed over the entire season, is lowest for the JHUAPL-DMD model forecasts, at both the national and regional level (Table 3). In addition, the JHUAPL-DMD model has lower WIS and MAE than the AI4Casting_Hub-Quantile_Baseline model, at both the national and regional level. VTSanghani-PRIME has lower MAE than the baseline model, at both the national and regional level, achieving similar performance to JHUAPL-DMD model on national MAE (Table 3). The UBCStat-Climate model has higher than baseline MAE and WIS, at both the national regional level.

**Table 3.** National and regional performance of UBCStat-Climate, VTSanghani-PRIME, AI4Casting_Hub-Quantile_Baseline, and JHUAPL-DMD across 2024-2025 Canadian RSV season. Model with best performance in each column is bolded. All models with performance better than AI4Casting_Hub-Quantile_Baseline are underlined.

| Model | Canada WIS (↓) | Region WIS (↓) | Canada MAE (↓) | Region MAE (↓) |
| --- | --- | --- | --- | --- |
| UBCStat-Climate | 1.22 | 1.24 | 2.17 | 2.34 |
| VTsanghani-PRIME | 0.73 | 1.43 | **0.58** | <u>1.73</u> |
| AI4Casting_Hub-Quantile_Baseline | 0.54 | 1.03 | 1.15 | 1.85 |
| JHUAPL-DMD | **0.37** | **0.68** | **0.57** | **1.09** |

Unlike the U.S. and Canadian 2024-2025 influenza and RSV seasons, which exhibited a clear peak (or peaks), the Canadian 2024-2025 COVID-19 season consisted of primarily a decrease, with positive percent of laboratory detections falling from ≈20% to ≈2%, before rising slightly to ≈4% (Fig. 8A).

**Figure 8.**
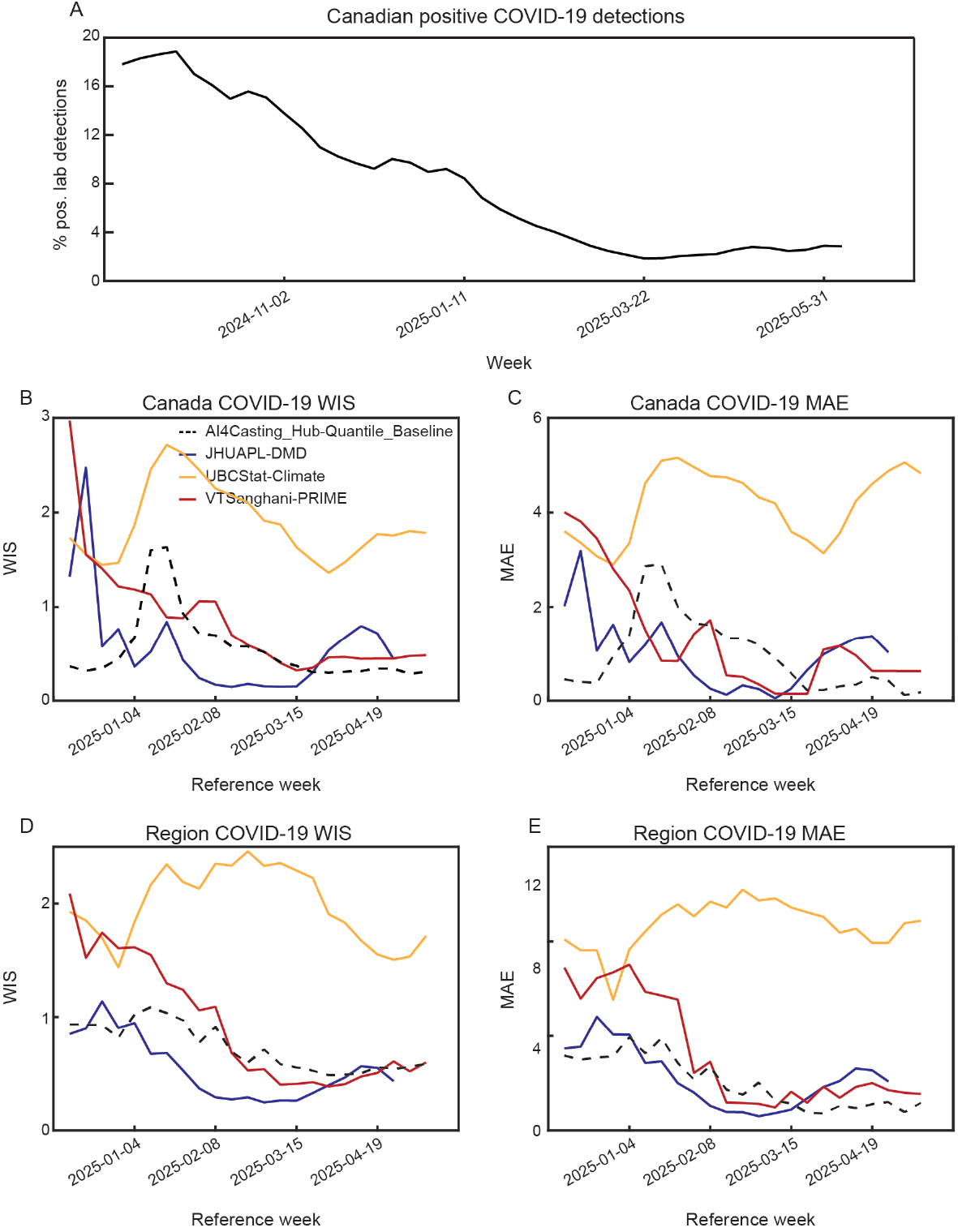
Performance of JHUAPL-DMD model forecasting Canada percentage of positive laboratory COVID-19 detections, by submission week. (A) Percent of positive COVID-19 laboratory detections in Canada over the 2024-2025 forecast season. (B)–(C) National WIS and MAE, across submission weeks, for JHUAPL-DMD, UBCStat-Climate, VTSanghani-PRIME, and AI4Casting_Hub-Quantile_Baseline. (D)–(E) State WIS and MAE, across submission weeks, for JHUAPL-DMD, UBCStat-Climate, VTSanghani-PRIME, and AI4Casting_Hub-Quantile_Baseline.

During the decay of COVID-19, we find that the JHUAPL-DMD model achieves the best WIS and MAE, at both the national and regional level, compared to all other models (Fig. 8B–D – reference week up to 2025-03-15). However, during the slow rise in COVID-19 during the spring, we find that JHUAPL-DMD’s performance decreases, with MAE and WIS rising above the the AI4Casting_Hub-Quantile_Baseline model (Fig. 8B–D – reference week after 2025-03-15). Examining an individual national forecast trajectory during this period of increasing COVID-19, we see that the JHUAPL-DMD model continues to forecast decreasing percent positive laboratory detections (Fig. 6, right column bottom row – blue line).

Evaluating the models on their median WIS and MAE for COVID-19, across the entire season, we find the AI4Casting_Hub-Quantile_Baseline model is the best performing model, with the lowest national WIS and MAE, and a regional MAE that is very similar to the JHUAPL-DMD model (Table 4). The JHUAPL-DMD model has the lowest regional WIS (Table 4). We note again that, the performance of the JHUAPL-DMD model is computed including the two submission weeks with erroneous forecasts. We suspect that, given the general trends we see on the other submitted weeks (Fig. 6), this may be the source of the lower performance of the JHUAPL-DMD model, relative to the baseline model. The VTSanghani-PRIME model outperforms the baseline model only on regional WIS, and the UBCStat-Climate model has higher MAE and WIS, at both the national and regional level.

**Table 4.**
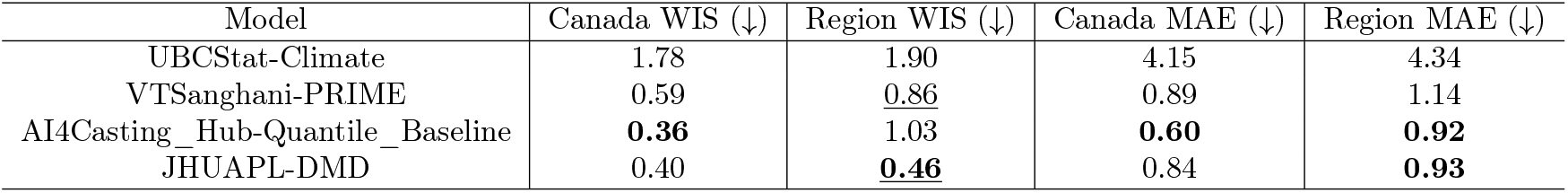
National and regional performance of UBCStat-Climate, VTSanghani-PRIME, AI4Casting_Hub-Quantile_Baseline, and JHUAPL-DMD across 2024-2025 Canadian COVID-19 season. Model with best performance in each column is bolded. All models with performance better than AI4Casting_Hub-Quantile_Baseline are underlined.

Given the improvement in performance we found when leveraging DMD with memory on the FluSight Forecast Hub data, we asked whether we may additionally find a benefit when applying DMD with memory to the AI4CastingHub data. Importantly, given the differences between the data, a similar improvement may not be *a priori* assumed.

For all three diseases, we find that – during the respective peaks (or rise, in the case of COVID-19) – DMD with memory is able to reduce the national and regional MAE (Fig. 9A–B, 10A–B, 11A–B). Individual forecast trajectories from the JHUAPL-DMD_mem model illustrate the ability of this approach to not only avoid overestimating the peak magnitudes, but also to capture the associated dynamics (Figs. 9C, 10C, 11C). Integrating the DMD with memory model forecasts during the peak weeks, with the sliding DMD model forecasts during the pre- and post-peak weeks, leads to improved regional and national MAE (Tables 5–7), with the exception of regional RSV WIS and MAE (Table 6). In the case of the RVDSS data, we do not find that thresholding impacts the median MAE and WIS significantly (Tables 5–7). This is likely due both to AI4CastingHub using the median to assess season performance, as compared to the FluSight Forecasting Hub that used the mean (Sec. 2.7), and because the JHUAPL-DMD model forecasts for RVDSS did not exhibit as large an overestimation of the peak magnitudes as they did in the U.S. influenza forecasts. Collectively, our analysis of the JHUAPL-DMD model’s performance on AICastingHub’s RVDSS data provides further evidence for the strengths of sliding DMD, particularly in the context of new data sets that have not been extensively analyzed before, and illustrates the fact that DMD with memory [31] may be a general way to improve performance during the season peak.

**Table 5.** National and regional performance of JHUAPL-DMD models across 2024-2025 Canadian influenza season. Model with best performance in each column is bolded. Note that these results are computed **not** including the two weeks JHUAPL-DMD submissions were affected by erroneous data loading.

| Model | Canada WIS (↓) | Region WIS (↓) | Canada MAE (↓) | Region MAE (↓) |
| --- | --- | --- | --- | --- |
| JHUAPL-DMD | 1.50 | 1.82 | 2.38 | 3.40 |
| JHUAPL-DMD threshold | 1.49 | 1.81 | 2.38 | 3.40 |
| JHUAPL-DMD_mem | **1.09** | **1.78** | **1.66** | **3.37** |

**Table 6.** National and regional performance of JHUAPL-DMD models across 2024-2025 Canadian RSV season. Model with best performance in each column is bolded. Note that these results are computed **not** including the two weeks JHUAPL-DMD submissions were affected by erroneous data loading.

| Model | Canada WIS (↓) | Region WIS (↓) | Canada MAE (↓) | Region MAE (↓) |
| --- | --- | --- | --- | --- |
| JHUAPL-DMD | 0.36 | **0.68** | 0.53 | **1.10** |
| JHUAPL-DMD threshold | 0.36 | **0.68** | 0.53 | **1.10** |
| JHUAPL-DMD_mem | **0.28** | **0.68** | **0.39** | 1.17 |

**Table 7.** National and regional performance of JHUAPL-DMD models across 2024-2025 Canadian COVID-19 season. Model with best performance in each column is bolded. Note that these results are computed **not** including the two weeks JHUAPL-DMD submissions were affected by erroneous data loading.

| Model | Canada WIS (↓) | Region WIS (↓) | Canada MAE (↓) | Region MAE (↓) |
| --- | --- | --- | --- | --- |
| JHUAPL-DMD | 0.38 | 0.42 | 0.84 | 0.85 |
| JHUAPL-DMD threshold | 0.38 | 0.42 | 0.84 | 0.85 |
| JHUAPL-DMD_mem | **0.24** | **0.38** | **0.40** | **0.68** |

**Figure 9.**
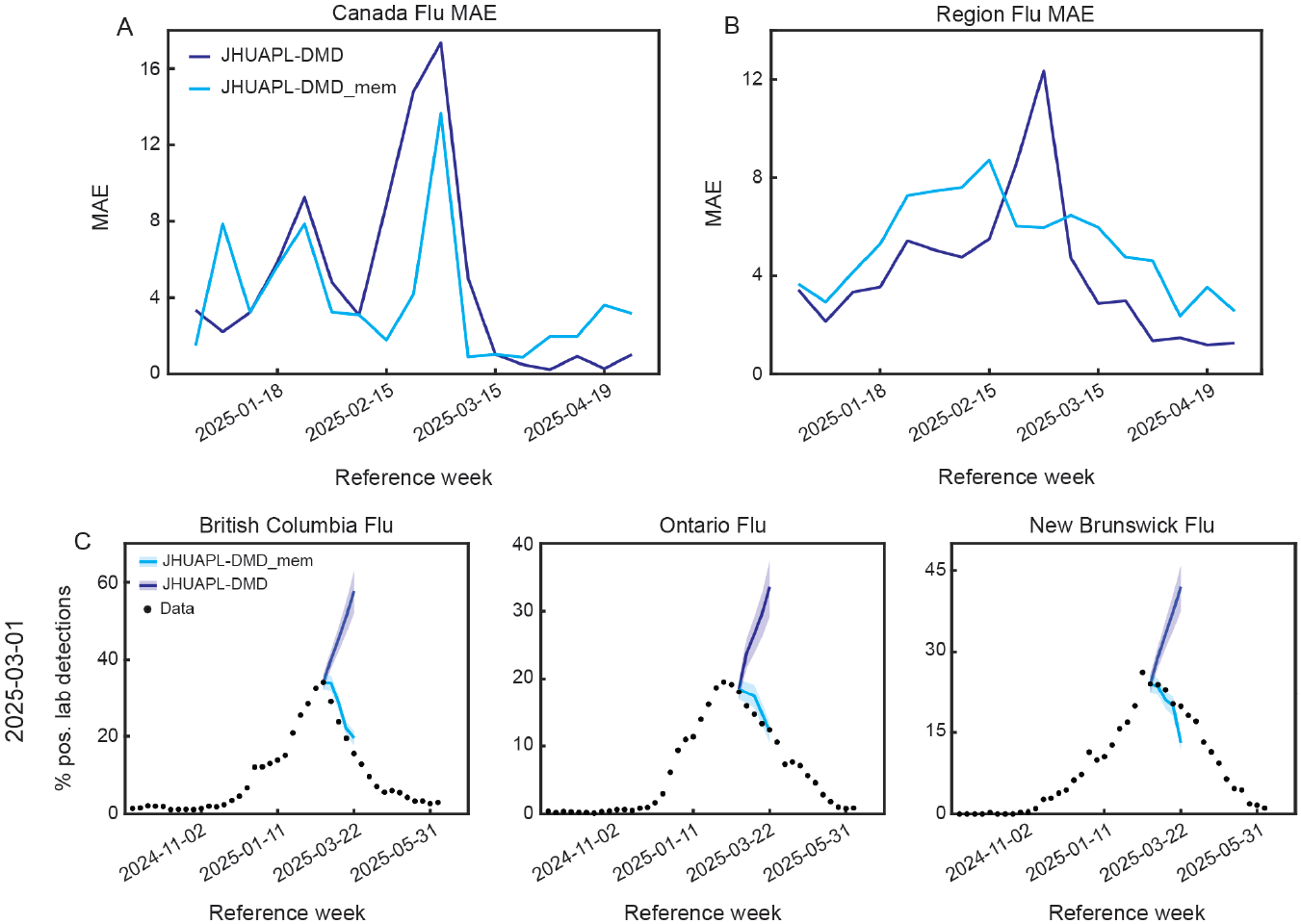
Utiliziation of memory improves the performance of JHUAPL-DMD forecasting during the Canadian influenza season’s peaks. (A)–(B) National and regional MAE, across submission weeks, for JHUAPL-DMD and JHUAPL-DMD_mem. (C) Example forecasts for British Columbia, Ontario, and New Brunswick percent positive influenza laboratory detections by JHUAPL-DMD and JHUAPL-DMD_mem models.

**Figure 10.**
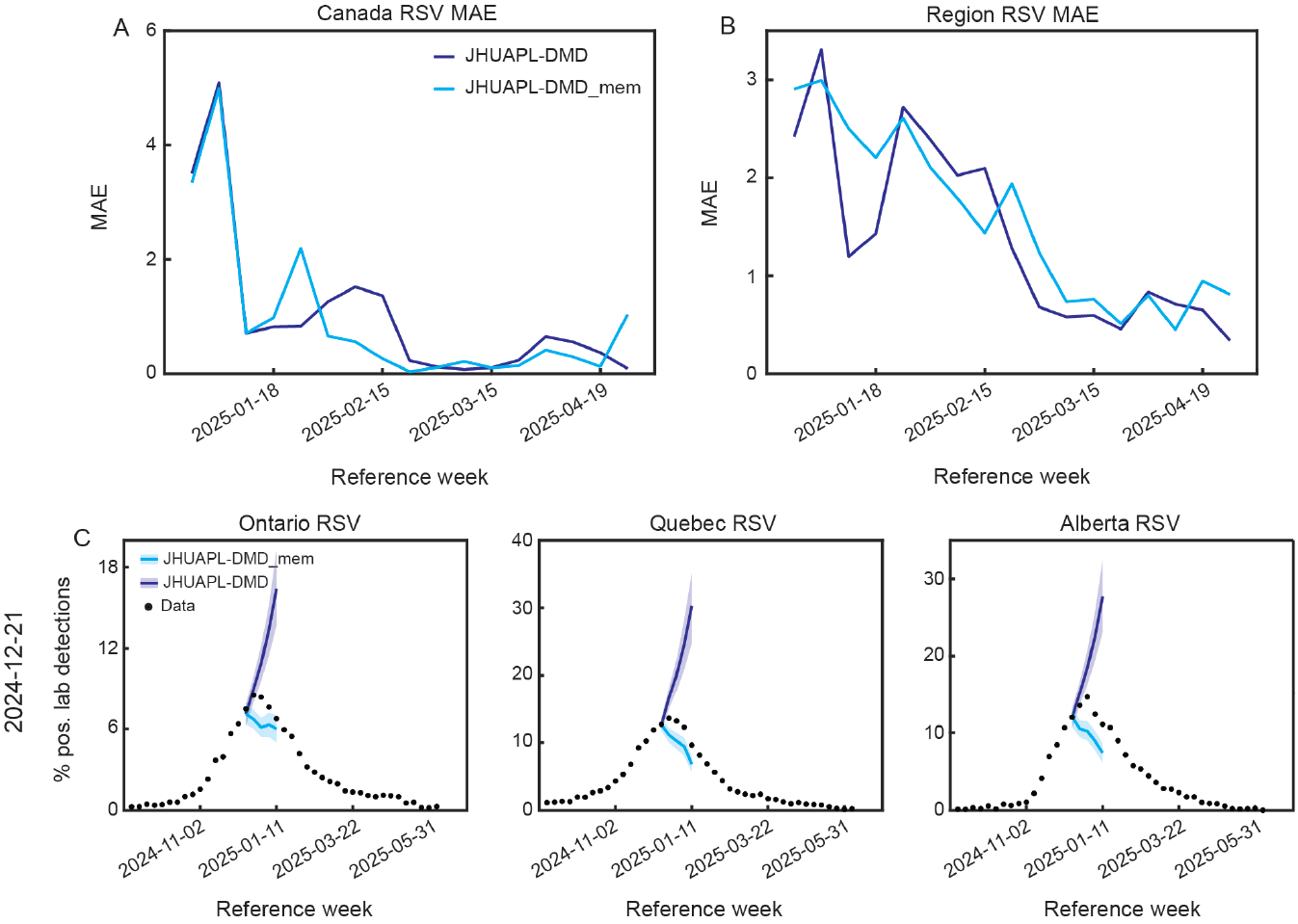
Utilization of memory improves the performance of JHUAPL-DMD forecasting during the Canadian RSV season’s peak. (A)–(B) National and regional MAE, across submission weeks, for JHUAPL-DMD and JHUAPL-DMD_mem. (C) Example forecasts for Ontario, Quebec, and Alberta percent positive RSV laboratory detections by JHUAPL-DMD and JHUAPL-DMD_mem models.

**Figure 11.**
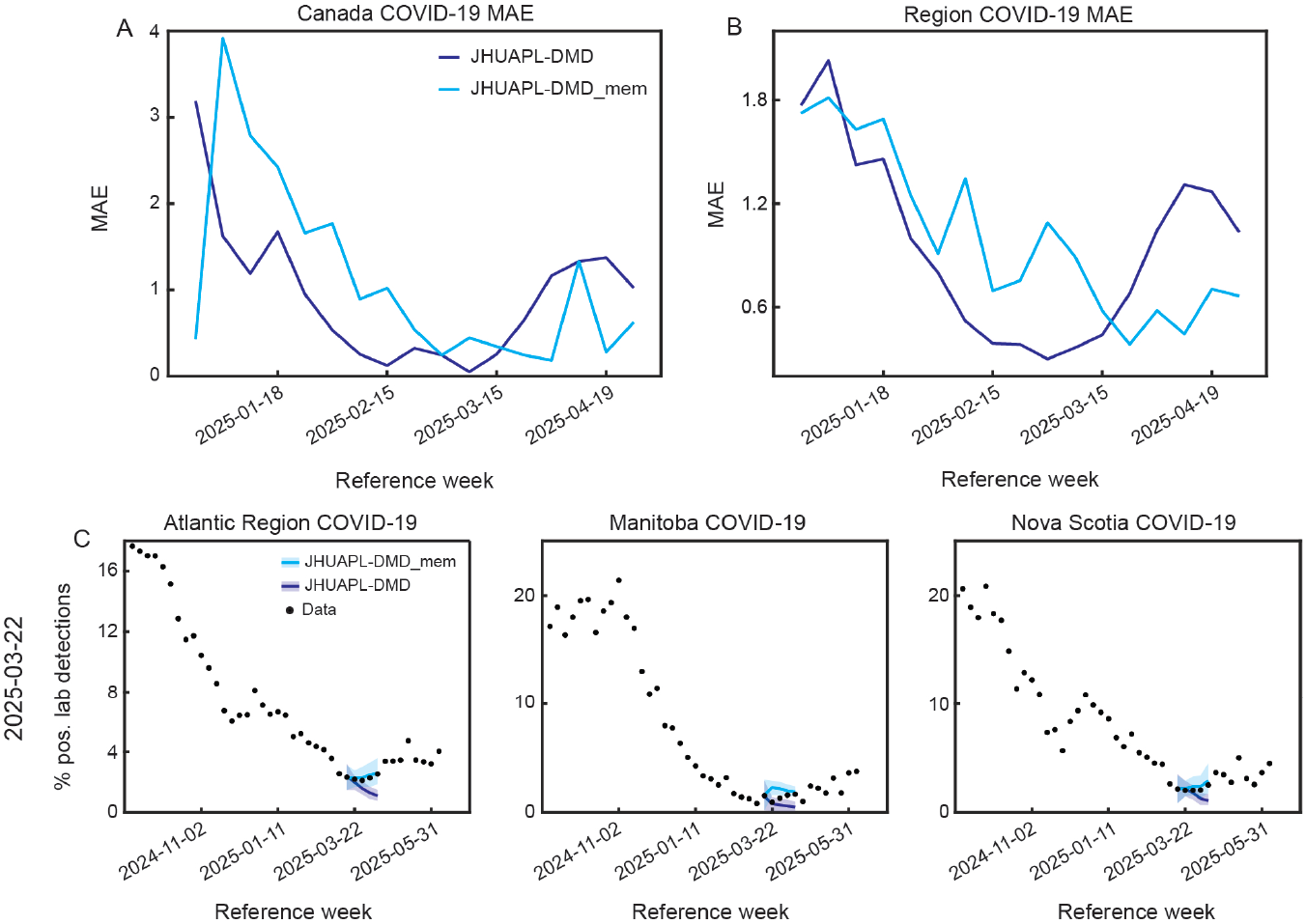
Utiliziation of memory improves the performance of JHUAPL-DMD forecasting during the Canadian COVID-19 season’s rise. (A)–(B) National and regional MAE, across submission weeks, for JHUAPL-DMD and JHUAPL-DMD_mem. (C) Example forecasts for Atlantic Region, Manitoba, and Nova Scoatia percent positive COVID-19 laboratory detections by JHUAPL-DMD and JHUAPL-DMD_mem models.

## 4 Discussion

Motivated by the need for light-weight, data-driven forecasting methods, we benchmarked sliding Hankel dynamic mode decomposition (DMD) [20, 21], an efficient and interpretable algorithm with theoretical dynamical systems grounding [20], on predicting influenza, RSV, and COVID-19 indicators in the U.S. and Canada. We found that sliding DMD robustly achieves similar or better performance to other models during the initial increase and subsequent decrease of targeted outcomes, illustrating the ability of temporally-local DMD models to capture dynamical trends. However, outside these periods (i.e. during the peak of the season), when the dynamics transition across dynamical regimes, sliding DMD’s lack of long-term temporal information leads to significant overshooting (or undershooting, in the case of COVID-19). Integration of DMD with memory [31], which equips sliding DMD with the ability to selectively leverage relevant past information to better inform its prediction, improves performance around the seasons’ peaks. This observation, however, results from retrospective analysis and required concurrent use of thresholding forecasts that were too large, suggesting that more innovation in DMD with memory is needed to make the method more robust.

We note that, to our knowledge, our work is the first to systematically study the performance of Koopman operator theoretic methods on live forecasting data. While benchmarking on static datasets is an important step, and has been done in some work combining Koopman operators and machine learning [44], having access to all the data can lead to “over-fitting”, even when cross validation is used, as hyper-parameters can be tuned until the best performance is found. For non-stationary data, where the optimal model for the known data might not be optimal for data coming from yet unseen dynamical regimes, this extra tuning may be problematic. We hope this work inspires others in the Koopman operator theory community to benchmark their models on live data, such as the forecasting Hubs we utilized.

The efficiency of sliding DMD was apparent when generating forecasts. Thousands of trajectories could be sampled within minutes to (at most) a couple of hours on a personal laptop computer. This was despite the fact that the code was not fully optimized. The low demand on compute further illustrates the potential Koopman operator theoretic methods have for enabling local (e.g., state, province, county, region) health departments to model their own data, even if they have limited computational resources. We believe this is an important direction that should continue to be embraced by the data-driven dynamics community.

### 4.1 Limitations and future directions

Our work has several important limitations that should be addressed in future work. First, our method for generating probablistic forecasts (Appendix B) utilizes uncertainty as computed across the entire season, instead of uncertainty that is more local in time. In addition, the exact approach for determining this uncertainty was done by heuristically fitting the week-to-week differences to a Gaussian. This led to less control of the quantiled forecasts and may contribute to the larger gap between the JHUAPL-DMD_mem threshold and CMU-TimeSeries model WIS values (Table 1), as compared to the MAE values. Better use of statistical methods to compute the uncertainty and integrate it with the sliding DMD forecasting is needed.

Second, while we retrospectively found improvement in forecast quality when using DMD with memory, this required identifying when to deploy the DMD with memory model and what the best hyper-parameters were. Both of these could be guided by more principled methods, which future work should explore. Our forecasts make use only of time-delay observables. While powerful, time-delays are not the only kind of lifting that is possible to use. In general, there is a need in the Koopman operator theory community to develop more theory for non-stationary dynamical systems [36]. One possible approach to optimization might include permuting over or sampling from a large number of parameter and lifting function combinations, and using ML approaches to learn the relationships between inputs (i.e. time-series), characteristics (history, length, time-frequency, observables, seasonality), hyper-parameters, and optimal performance of sliding DMD with memory. This learned model could then be leveraged to automatically preselect the optimal set of parameters and lifting functions, for a given input.

Third, our forecasts came from a single sliding DMD model. Forecasts ensembled from multiple models can be more robust and creating a family of DMD models to ensemble over could provide better performance. We expect that using DMD models built from a data across a range of time-scales (e.g., multi-resolution DMD [45]) would be especially powerful. Indeed, relevant long-range dynamical information has been shown to be extractable from global DMD models [46] in the context of climate science. The extent to which this holds for infectious diseases remains to be seen.

Finally, while our work focused on the quality of forecasts generated by sliding DMD, another appealing aspect of Koopman operator theoretic methods is the ability to learn models with interpretable spatial information. Analyzing the extent to which this information can provide insight on the spatial dynamics of influenza, RSV, and COVID-19 would be insightful. This kind of information is not easily accessible in blackbox ML and statistical methods, and could be useful for public health decision makers.

## Data Availability

All data used in our analysis is publicly available:

- CDC FluSight– https://github.com/cdcepi/FluSight-forecast-hub
- AI4CastingHub RVDSS– https://github.com/ai4castinghub/rvdss-forecast

All code used in our analysis will be made publicly available upon acceptance.

## Acknowledgments

We thank Dean Huang and Jordan Garret for feedback on presentation of Fig. 1. We thank Igor Mezić and Maria Fonoberova for helpful discussions on DMD with memory. We thank Sarabeth Mathis and Rebecca Borchering for their assistance with our CDC FluSight submissions and Siddhesh Kadam for his assistance with our AI4CastingHub submissions.

## A DMD with memory

To enable our sliding DMD method to have access to longer time-scale information, beyond onyl what was in the sliding window, we used DMD with memory [31]. This approach looks for windows in the past whose associated Koopman eigenvalues and modes are “similar” to those currently observed (Fig. 1D–G). In the case that such a match is found, the known dynamics that occurred previously are used for forecasting.

The first step in using DMD with memory is to generate a “data bank” with the Koopman eigenvalues and modes associated with previous windows. For the FluSight submissions, we used the historic data that is available, which contains hospitalizations from the past few seasons. For the AI4CastingHub submissions, we used the historic data that is available, which contains the percent of positive influenza and RSV laboratory tests for the past 10 years, and the percent of positive COVID-19 laboratory tests for the past few years. When constructing the sliding DMD model, all forecast targets are modeled together. This allows for capturing possible spatio-temporal dynamics between regions. However, because of the large number of regions, we expected that, when comparing the dynamics in the past with the current dynamics, the number of associated Koopman eigenvalues and the size of the modes may lead to poor quality matches. Therefore, we instead compute the Koopman eigenvalues and modes for each region separately.

Because there is little historic data available, we reasoned it may improve our ability to leverage DMD with memory if we compared a given region’s Koopman eigenvalues and modes with the saved Koopman eigenvalues and modes from other regions. However, the significant differences between forecasting targets (e.g., differences in population, density, healthcare) led us to expect that some regions may not provide good information about other regions. To address this, we set a threshold, *τ*_pop_ ∈ [0, 1], that determines the maximum relative difference in population that is allowed for one region to be compared to another region. For instance, setting *τ*_pop_ = 0.5 requires two regions to have a population ratio of greater than 50% in order for the Koopman eigenvalues and modes of one region to be compared to the other region. For the FluSight submissions, we set *τ*_pop_ = 0.75 and for the AI4CastingHub submissions, we set *τ*_pop_ = 0.25.

Finally, as noted in Sec. 2.3, in our implementation of we do not use only a single match, as was done previously [31]. Instead, we allow for multiple matches, and average their individual forecasts to get a (hopefully) more informative forecast. When implementing this, we found that when a large number of matches were found, including all of them could generate worse predictions then when just using the “best” matches. Therefore, we set a threshold, *τ*_match_ ∈ ℕ, which determined the maximum number of matches used to generate a DMD with memory forecast. For both FluSight and AI4CastingHub submissions, we set *τ*_match_ = 10. To determine which matches to keep, in the case that more than *τ*_match_ were found, we rank the matches based on the product of the Wasserstein and Euclidean distance. That is, we compute the Wasserstein distance between the Koopman eigenvalues of the current window and the previous time window, we compute the Euclidean distance between the Koopman modes of the current window and the previous time window, and then we multiply these together. We keep only the *τ*_match_ matches with the lowest product distance.

## B Quantile forecasting

To generate probabilistic forecasts, we sample noise, *δ*_*i*_(*t*) ∼ *𝒩* (0, *σ*_*i*_), for each forecasting target (e.g., state, region, nation) and add this to the observed data. To determine *σ*_*i*_, we compute the distribution of Δ_*i*_ = {|*x*_*i*_(*t*) − *x*_*i*_(*t* + 1) |,− |*x*_*i*_(*t*) − *x*_*i*_(*t* + 1) |}, for all past values of *x*_*i*_(*t*). In the case of the FluSight submissions, this was done on two seasons worth of historic data. Historic data was used for the first few weeks in generating AI4CastingHub submissions, but then we switched to only using the data that had been observed in the 2024-2025 season, as this led to less conservative confidence intervals. Inspection of Δ_*i*_ led to the identification of two distributions. One that is approximately normal, and one that is centered around 0. This second distribution comes from the relatively static part of the time-series at the end of the season. We mask out this first distribution by removing any values of Δ_*i*_ that are less than 0.5. A Gaussian was then fit to the remaining part of Δ_*i*_.

This is a heuristic approach and can certainly be improved upon. However, it is worth noting that despite this, the 50% coverage of our FluSight submissions was ≈ 52% and the 95% coverage was ≈ 86%.

In the case when DMD with memory is used, an initial thought was to implement the same approach to generate quantiled forecasting. However, we found that sampling noisy data, computing the associated Koopman eigenvalues and modes, and then looking for matches led to worse performance. We imagine that this is due to the fact that this noisy data is not actually representative of the data that is observed, and can lead to matches with very different dynamical behavior. Therefore, to generate quantiled forecasts we did the following. DMD with memory was applied to the true (non-noisy) data. In the case where no match was found, we used the standard sliding DMD forecast 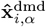, which includes the quantiles computed as noted above. In the case where matches are found, we get 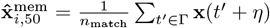 (Sec. 2.3). That is, the average behavior subsequent to the matched time-windows in the past is used as the 50^th^ quantile forecast. To get the other quantiles, we scale the forecasted quantiles from sliding DMD. That is, 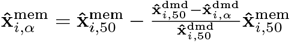. In this way, the DMD with memory quantiles are the same as sliding DMD quantiles, relative to the median forecast.

## Footnotes

1 https://4castinghub.uoguelph.ca/forecast-evaluation-dashboard/

2 https://reichlab.io/flusight-dashboard/eval.html

